# Causal analysis shows evidence of atopic dermatitis leading to an increase in vitamin D levels

**DOI:** 10.1101/2020.04.22.20075077

**Authors:** Daniel R Drodge, Ashley Budu-Aggrey, Lavinia Paternoster

## Abstract

Atopic dermatitis (AD) patients have been observed to have lower vitamin D levels. Previous studies have found little evidence that vitamin D levels causally influence the risk of AD, but the reverse direction has not yet been investigated.

Here we used Mendelian Randomization to assess the causal relationship between AD and serum vitamin D levels, using genetic data from the most recent GWA studies of vitamin D and AD.

There was little evidence for vitamin D levels causally influencing AD risk (odds per standard deviations increase in log-transformed vitamin D levels =1.233, 95% CI 0.927 to 1.639, *P*-value =0.150). However, genetic liability for AD raises serum vitamin D levels by 0.043 (95% CI 0.017 to 0.069) standard deviations per doubling of odds of disease (*P*-value =0.001). The AD-associated filaggrin (*FLG*) mutation R501X appears to show a particularly strong relationship with vitamin D. However, the relationship between AD and vitamin D holds when R501X is omitted (0.018, 95% CI 0.004 to 0.031, *P*-value =0.008).

We found evidence that AD is causally associated with an increase in serum vitamin D levels. Whilst the AD-associated *FLG* gene has a particularly strong relationship with vitamin D, other AD SNPs show a consistent direction of effect, suggesting that AD more generally influences serum vitamin D levels.

## Introduction

Atopic dermatitis (AD, eczema) is an inflammatory skin condition which typically presents as erythema, scaling and urticaria (Weidinger and Novak 2016). U.S. prevalence is of the order of 10%, higher in children than adults (Drucker et al. 2017). Moderate and severe disease imposes a functional and psychological burden on sufferers, as well as the risk of complications such as bacterial superinfection, eczema herpeticum and erythroderma. Treatments currently follow several strategies: restoration of barrier function through topical emollients, reduction of inflammation through immunomodulatory agents, and second-line treatments including phototherapy (Rodenbeck et al. 2016). A treatment of ongoing controversy is vitamin D supplementation - the most recent Cochrane review concluded no benefit and warned of toxic effects of high doses (Bath-Hextall et al. 2012), but a more recently-performed meta-analysis found a reduction in SCORAD in patients receiving supplementation (Hattangdi-Haridas et al. 2019).

AD and vitamin D have been linked in observational studies: sufferers have reduced levels of the hormone, more clearly so in children, and some evidence exists that deficiency is more marked in severe disease (Palmer 2015). Vitamin D plays a role in calcium and phosphate homeostasis, and its deficiency has also been implicated in a variety of immunological disease states (Rosen et al. 2012). It derives from cholecalciferol, a steroid converted from the cholesterol precursor 7-dehydrocholesterol in the skin, under the action of ultraviolet radiation. Hydroxylation in the liver produces 25-hydroxycholecalciferol (Vitamin D; 25-OHD), which the kidneys convert to an active form, 1,25-dihydroxycolecalciferol (1,25-OHD).

In questions of aetiology and causality, Mendelian Randomization (MR) can be of use. It allows causal inference to be drawn from population data, namely genome-wide association studies (GWAS) (Budu-Aggrey and Paternoster 2019). To assess the effect of an exposure on an outcome, MR uses exposure-associated single nucleotide polymorphisms (SNPs) as a proxy for the exposure of interest. These mutations are randomised at conception, thus limiting the influence of confounders on the allocation of exposure. MR is analogous to a randomised controlled trial without the cost and ethical difficulty of assigning disease states or known toxic environmental factors as exposures. The key underlying assumption is that each SNP we use in this way acts on the outcome only through its upstream action on the exposure – this is violated if it affects it, whether directly or through an alternative pathway.

Manousaki and colleagues previously used MR to ascertain whether there was any evidence that vitamin D levels causally influenced AD, using a two sample MR method, with 25-OHD levels GWAS data from the SUNLIGHT consortium, and the AD GWAS data from the EAGLE consortium (Manousaki et al. 2017). Here we extend this analysis using a recently published larger vitamin D GWAS meta-analysis, which includes 401,460 UK Biobank participants (Manousaki et al. 2020). We also include an investigation of the inverse causal direction, i.e. whether there is any evidence that genetic risk of AD causally influences vitamin D.

## Results and Discussion

MR analyses of AD and 25-OHD are shown in Table 1: change in serum 25-OHD level is presented log-transformed, normalised to zero mean with unit standard deviation, consistent with the units used by Manousaki. There was little evidence for vitamin D levels causally influencing the risk of AD (odds ratio per SD change in log-transformed 25-OHD levels = 1.233, 95% CI 0.927 to 1.639, *P*-value =0.150), a finding in keeping with the 2017 analysis which found an effect size of 1.12, (95% CI 0.92 to1.37, *P*-value =0.270) (Manousaki et al. 2017). However, we did find evidence for AD causally increasing vitamin D levels – an effect of 0.043 SD increase in log-transformed 25-OHD concentration per doubling odds of AD (95% CI 0.017 to 0.069, *P*-value =0.001).

**Table 1.** Summary of Mendelian Randomization analyses performed for forward and reverse causation between vitamin D (25-OHD) and genetic risk of Atopic Dermatitis (AD). Single nucleotide polymorphism (SNP); Standard Deviation (SD); 95% confidence interval (95% CI).

| Exposure | Outcome | N SNPs | Effect (95% CI) | $P$-value | Units |
| --- | --- | --- | --- | --- | --- |
| 25-OHD | AD | 68 | 1.233<br>(0.927 to 1.639) | 0.150 | Odds per SD change in log-transformed 25-OHD levels |
| AD | 25-OHD | 24 | 0.043<br>(0.017 to 0.069) | $1.33 \times 10^{-3}$ | SD change in log-transformed 25-OHD levels per doubling odds of AD |

Tests of validity of this analysis were performed, which found strong evidence of heterogeneity (Cochran Q =282, *P*-value < 2.2×10^−16^) and horizontal pleiotropy (Egger intercept = -0.012; 95% CI= -0.018 to -0.006; *P*-value=5.4×10^−4^) (Figure S1). A substantial contribution to this appeared to be the particularly strong result from one SNP, R501X (rs61816761), a functional mutation in the *FLG* region (Figure S2). Other functional *FLG* mutations are known and so we further examined the association between the 4 most common of these (R501X[rs61816761], 2282del4 [rs41370446], R2447X [rs138726443], and S3247X [rs150597413]) and vitamin D from the GWAS. All except 2282del4 were available in the vitamin D GWAS and found to be strongly associated with 25-OHD levels (Figure S3). To investigate whether the causal relationship was driven entirely by the *FLG* region we repeated the MR omitting this locus. The effect estimate was smaller, but still gave evidence of causality: 0.018 (0.004 to 0.031, *P*-value= 0.008) (Table 1, Figure 1). Sensitivity analyses found less evidence of heterogeneity when excluding the *FLG* locus from the AD instrument (Cochran’s Q=60, *P*-value=2.3×10^−5^) and little evidence of horizontal pleiotropy (Egger intercept = 0.001; 95% CI= -0.005 to 0.006; *P*-value=0.782) (Figure S4).

**Figure 1.**
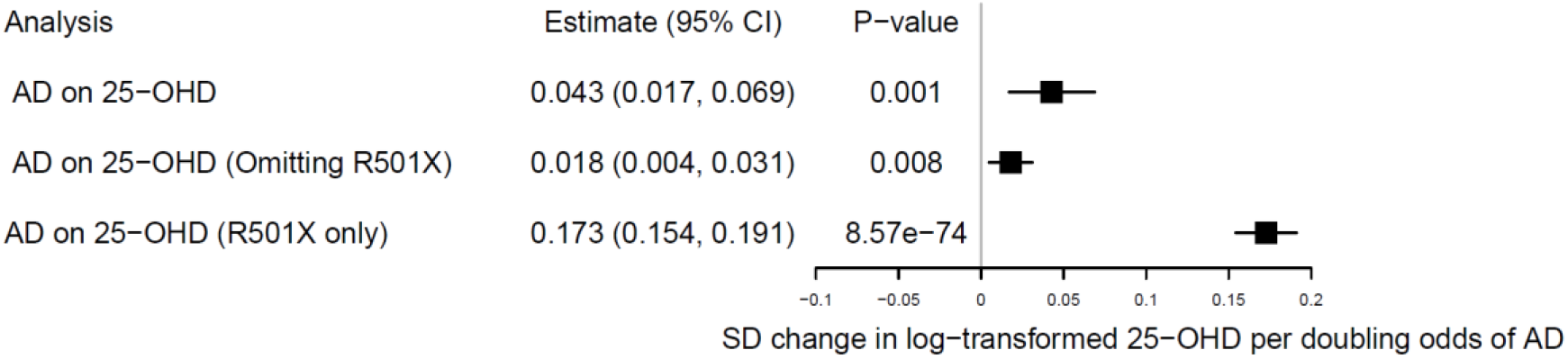
MR causal estimates of the effect of genetic risk of atopic dermatitis (AD) on vitamin D (25-OHD), using (i) an AD instrument comprising all 24 GWAS SNPs, (ii) an AD instrument excluding the *FLG* R501X functional mutation and (iii) an AD instrument of the FLG R501X functional mutation alone. Standard Deviation (SD)

To explore whether this causal relationship is limited to AD, or whether inflammatory disease per se might confer any change in vitamin D levels, additional MR analyses were carried out for other inflammatory disease for which there were strong instruments available (Figure 2), including inflammatory skin condition, psoriasis. There was little evidence that other inflammatory diseases influenced 25-OHD levels. For type 1 diabetes mellitus there was some evidence of a causal decrease in vitamin D (-0.004 standard deviations per doubling odds of disease [95% CI = -0.007 to 0.000], *P*-value = 0.044). There was also some evidence of a causal increase in vitamin D levels with systemic lupus erythematosus (0.003 standard deviations per doubling odds of disease [95% CI = 0.000, 0.005], *P*-value = 0.017). These estimates might warrant further investigation, however the evidence is weak considering the number of diseases tested (i.e. a Bonferroni-corrected alpha for global alpha<0.05 is 0.005).

**Figure 2.**
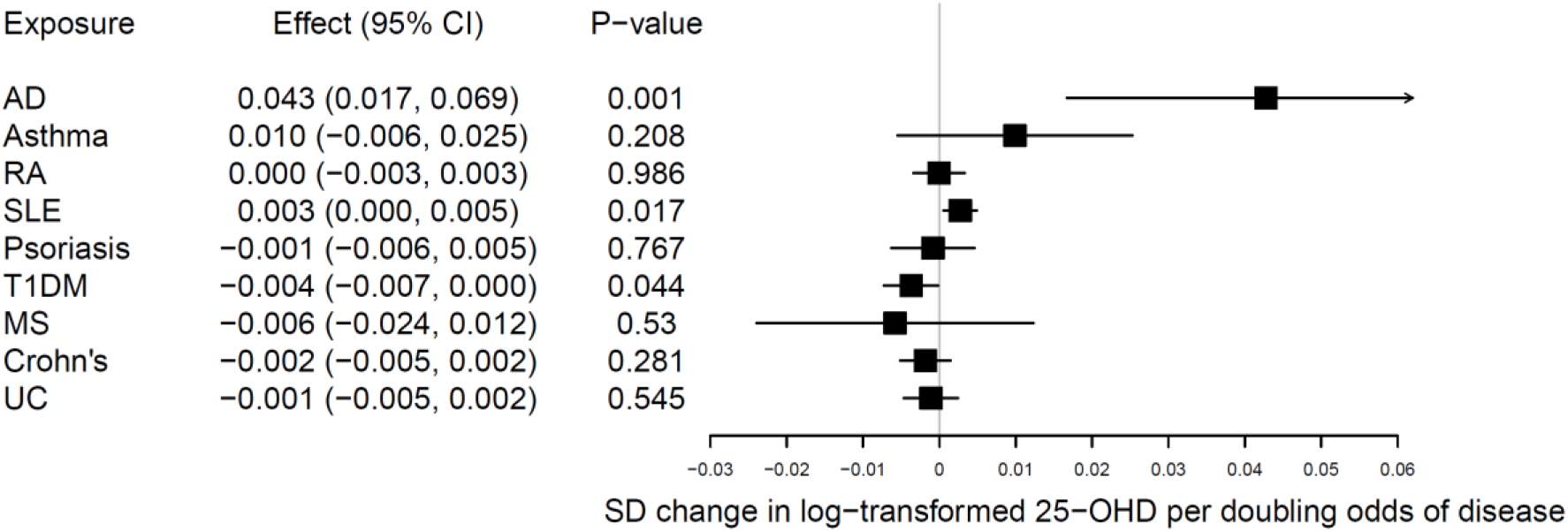
Causal effect estimates of various exposure disease states on serum 25-OHD expressed as standard deviation (SD) change in log 25-OHD per doubling odds of disease, as estimated using Mendelian Randomization. AD: atopic dermatitis; RA: rheumatoid arthritis; SLE: systemic lupus erythematosus; T1DM: type 1 diabetes mellitus; MS: multiple sclerosis; UC: ulcerative colitis.

## Discussion

We found evidence that (i) AD causes an increase in Vitamin D levels, (ii) although the filaggrin locus has a strong influence on this, a causal effect is still seen when omitting this locus; and (iii) no similar magnitude and direction of effect is seen in other atopic and autoinflammatory diseases.

We confirm that there is little evidence that vitamin D causally influences AD, as has previously been reported (Manousaki et al. 2017), but find evidence, from the reverse direction Mendelian Randomization analysis, that genetic risk of AD causally increases vitamin D levels. This relationship is opposite to what has been reported observationally, where AD patients are seen to have lower vitamin D (Palmer 2015). It thus undermines the rationale for using vitamin D supplementation as a therapy for AD. When causal estimates are opposite to those reported observationally, it is likely that this is due to the confounding structure masking the true nature of the relationship in the observational estimates. Confounders that have opposing effects on vitamin D and AD, that might explain this phenomenon include obesity, SES, pollution and latitude (Mesquita et al. 2013). In any case, our current study provides further evidence that raising vitamin D is unlikely to be an effective treatment for AD.

Our analysis found a particularly strong relationship between *FLG* variants and increased vitamin D. Filaggrin is a 400 kDa protein, consisting of several repeating segments, which serves multiple functions in maintaining the skin barrier (Brown and Irwin McLean 2012). Several functional *FLG* mutations are known, the most common being *FLG* null mutation R501X - homozygotes almost all have skin disease (McLean 2016). Other common *FLG* null mutations include 2282del4 (rs41370446), R2447X (rs138726443), and S3247X (rs150597413). We were able to test the association between vitamin D and R501X, R2447X and S3247X and found >0.1SD change in 25-OHD per mutation at each of these sites. The fact that we observed substantial heterogeneity and evidence of horizontal pleiotropy when we included the *FLG* locus in our AD instrument (which reduced substantially when this locus was removed), suggests that *FLG* has a direct influence on vitamin D, that bypasses AD. A link between *FLG* and vitamin D has been previously reported in the same direction that we report here (Thyssen et al. 2012). They sought to explain this with a UVB-VitD3 hypthosis, whereby trans-urocanic acid (UCA), a breakdown product of filaggrin (generated when histidase metabolises histidine in filaggrin), provides epidermal protection against UVB (Mildner et al. 2010). Therefore, inheriting a *FLG*-null mutation (particularly one which reduces histidine in filaggrin – i.e. the R501X truncation reduces FLG from 413 to 35 histidine residues (Fauman 2020)) would result in increased UV absorption and increased vitamin D3 synthesis. It has been further proposed that such a mechanism might be advantageous in northern latitudes and is one explanation as to why a latitude-dependent gradient of *FLG* mutation frequency is observed (Thyssen and Elias 2017). Given, this strong relationship between *FLG* mutations and increased vitamin D, it would be sensible in future clinical trials of supplementation or phototherapy to investigate whether different effects are observed in AD patients with and without *FLG* mutations.

In addition to the *FLG*-vitamin D direct link, there also appears to be a general influence of AD on vitamin D, as evidenced by the causal effect observed after omitting the *FLG* locus from the AD instrument. It is unclear what the mechanism for this might be, whether there are other biological effects in the skin, similar to that seen for FLG, or whether there are behavioural mechanisms by which suffering from AD may in turn alter UV and/or vitamin D exposure. We also need to consider whether the vitamin D GWAS (from which our data were taken) included people with AD whose vitamin D status was raised because they were using vitamin D raising therapy. The study did adjust for vitamin D supplementation (Manousaki et al. 2020) and although phototherapy was not specifically commented on, the UK Biobank website showcase summary reports that only 69 of 410,256 subjects are coded as receiving some form of phototherapy (operative procedures OPCS4, skin, phototherapy S12) (UK Biobank) – if accurate and complete, this translates to a small proportion within the vitamin D GWAS and is unlikely to explain our findings.

In this study we confirm that there is little evidence that vitamin D causally influences AD. However, we did find evidence to suggest that the genetic risk of AD causally increases serum vitamin D levels, both through the direct effect of the FLG gene on vitamin D, and also through additional, likely more general, mechanisms. Our findings suggest that vitamin D supplementation as a treatment for AD is not supported, moreover that further investigation into the relationship between AD and vitamin D is warranted to determine the mechanisms of the reported causal relationship and suggest stratifying future vitamin D and phototherapy trials and epidemiological analyses by *FLG* status to be able to better evaluate the contribution of different mechanisms.

## Materials and Methods

### Genetic instruments and sources

To investigate the causal effect of genetically predicted vitamin D levels upon AD risk, a genetic instrument for 25-OHD levels was derived from SNPs reported to be most strongly associated (*P*-value < 5×10^−8^) in the most current GWA study by Manousaki and colleagues (N=443,734) (Manousaki et al. 2020) (Table S1). SNP-association estimates were presented for standardized log-transformed 25-OHD levels. A total of 59 independent SNPs (r^2^ threshold = 0.001) were identified using the TwoSampleMR R package clumping tool (Hemani et al. 2018) (Table S2). Full summary data was also made available by the authors to allow for MR analyses with vitamin D as the outcome.

To investigate the causal effect of AD genetic liability upon vitamin D levels, an AD genetic instrument was derived from the most current GWAS by the EAGLE consortium (Paternoster et al. 2015), where 24 loci were reported to be most strongly associated with AD (*P*<5×10^−8^) (Table S1 & S3). The index SNP from each GWAS locus was selected (except for 1q21.3, where the known functional variant rs6181671 replaced the index GWAS SNP rs61813875). Effect estimates were available for 18,900 cases and 84,166 controls of white European ancestry. AD GWAS summary statistics (with 23andMe participants excluded) was also made available (10,788 cases, 30,047 controls).

The genetic liability of additional inflammatory diseases upon vitamin D levels was also investigated. In doing so, published GWAS data was used to derive genetic instruments for asthma (Demenais et al. 2018), rheumatoid arthritis (Okada et al. 2014), systemic lupus erythematosus (Bentham et al. 2015), psoriasis (Tsoi et al. 2017), type 1 diabetes mellitus (Onengut-Gumuscu et al. 2015), multiple sclerosis (Beecham et al. 2013), Crohn’s Disease and ulcerative colitis (de Lange et al. 2017) using SNPs reported to be most strongly associated (*P*-value < 5.0×10^−8^) (Table S1; S4 to S11). SNP estimates were derived from individuals of white European ancestry. Given the overlapping risk variants for AD and asthma, the asthma instrument was filtered to exclude SNPs with nominal association with AD (p<0.05).

### Statistical Analyses

Two-sample MR was performed to investigate the casual relationship between AD and genetically predicted vitamin D levels using the TwoSampleMR package in R. Our genetic instrument for vitamin D levels was used to assess the causal effect upon AD risk using summary GWAS data from the most recent AD GWAS meta-analysis (Paternoster et al. 2015). Likewise, in the reverse direction, the AD genetic instrument defined was used to estimate the causal effect of AD genetic risk upon vitamin D levels (SD change in log 25-OHD levels) using summary GWAS data from the recent vitamin D GWAS (Manousaki et al. 2020). This was also performed with the additional inflammatory instruments created to estimate the causal effect of each trait upon vitamin D levels. As the GWAS data for AD and inflammatory traits did not include UK Biobank participants, there was no sample overlap in the MR analyses performed.

A causal estimate was obtained by applying the inverse-variance weighting (IVW) method, where the SNP-exposure and SNP-outcome associations were combined in a meta-analysis assuming multiplicative random effects. When presenting the causal effect upon vitamin D levels, causal estimates were multiplied by 0.693 to represent the SD change in log 25-OHD levels per doubling odds of AD (or inflammatory trait) genetic risk, as previously demonstrated (Gage et al. 2017). MR-Egger regression, weighted median analysis and the weighted mode-based estimate (MBE) were performed to investigate potential horizontal pleiotropy. SNPs found to influence the exposure and the outcome through different pathways could bias the causal estimate and violate the MR assumption that genetic instruments only have an effect on the outcome via the exposure investigated (Budu-Aggrey and Paternoster 2019). MR-Egger regression provides a valid causal estimate under the ‘InSIDE’ assumption, where each SNP-exposure effect is uncorrelated with the horizontal pleiotropic effect of the SNP. The intercept also allows the size of a pleiotropic effect to be determined (Bowden et al. 2015). The weighted median method provides a valid causal estimate if at least 50% of the information in the MR analysis comes from valid instruments (Bowden et al. 2016), and the weighted MBE provides a valid causal estimate assuming that the most frequent pleiotropy value is zero across the genetic instruments (Hartwig et al. 2017). We also characterised heterogeneity using Cochran’s Q statistic, which can indicate invalid instruments that are present due to the occurrence of pleiotropy (Bowden et al. 2018).

All analyses were carried out in R (www.r-project.org) unless otherwise stated. The code used to carry out these analyses is available on GitHub (https://github.com/abudu-aggrey/Atopic_Dermatitis_25OHD_MR).

## Data Availability

Datasets related to this article can be found at https://github.com/abudu-aggrey/Atopic_Dermatitis_25OHD_MR. Summary GWAs data for vitamin D (25-OHD) levels was made available by the authors upon request (Manousaki et al. 2020).

https://github.com/abudu-aggrey/Atopic_Dermatitis_25OHD_MR

## Conflicts of interest

LP has received personal fees from Merck for Scientific Input Engagement related to MR methodology.

## Acknowledgements

DD performed this research as part of the academic foundation training programme organised by Health Education England South West, and thanks Prof. D. Gunnell for mentoring support. DD, LP and AB-A work in a research unit funded by the UK Medical Research Council (MC_UU_00011/1). LP received funding from the British Skin Foundation (8010 Innovative Project) and the Academy of Medical Sciences Springboard Award, which is supported by the Wellcome Trust, The Government Department for Business, Energy and Industrial Strategy, the Global Challenges Research Fund and the British Heart Foundation [SBF003\1094].

We are grateful to the authors of the published GWAS studies for making their summary statistics available to allow this work to be possible.

